# Safety and immunogenicity of a recombinant tandem-repeat dimeric RBD protein vaccine against COVID-19 in adults: pooled analysis of two randomized, double-blind, placebo-controlled, phase 1 and 2 trials

**DOI:** 10.1101/2020.12.20.20248602

**Authors:** Shilong Yang, Yan Li, Lianpan Dai, Jianfeng Wang, Peng He, Changgui Li, Xin Fang, Chenfei Wang, Xiang Zhao, Enqi Huang, Changwei Wu, Zaixin Zhong, Fengze Wang, Xiaomin Duan, Siyu Tian, Lili Wu, Yan Liu, Yi Luo, Zhihai Chen, Fangjun Li, Junhua Li, Xian Yu, Hong Ren, Lihong Liu, Shufang Meng, Jinghua Yan, Zhongyu Hu, Lidong Gao, George F. Gao

## Abstract

**Background:** A safe and effective coronavirus disease 2019 (COVID-19) vaccine is urgently needed to control the ongoing pandemic. Although progress has been made recently with several candidates reporting positive efficacy results, COVID-19 vaccines developed so far cannot meet the global vaccine demand. We developed a protein subunit vaccine against COVID-19, using dimeric form of receptor-binding domain (RBD) as the antigen. We aimed to assess the safety and immunogenicity of this vaccine in humans and determine the appropriate dose and schedule for an efficacy study.

**Methods:** We did two randomized, double-blind, placebo-controlled, phase 1 and 2 trials for an RBD-based protein subunit vaccine, ZF2001. In phase 1 study, 50 healthy adults aged 18-59 years were enrolled and randomly allocated to three groups to receive three doses of vaccine (25 μg or 50 μg RBD-dimer, with adjuvant) or placebo (adjuvant-only) intramuscularly, 30 days apart. In phase 2 study, 900 healthy adults aged 18-59 years were enrolled and randomly allocated to six groups to receive vaccine (25 μg or 50 μg RBD-dimer, with adjuvant) or placebo (adjuvant-only) intramuscularly, with the former 3 groups given two doses and the latter 3 groups given three doses, 30 days apart. For phase 1 trial, the primary outcome was safety, as measured by the occurrence of adverse events and serious adverse events. The secondary outcome was immunogenicity as measured by the seroconversion rate and magnitude of antigen-binding antibodies, neutralizing antibodies and T-cell cytokine production. For phase 2 trial, the primary outcome included both safety and immunogenicity. These trials are registered with ClinicaTrials.gov, NCT04445194 and NCT04466085.

**Findings:** Between June 22 and September 15, 2020, 50 participants were enrolled to the phase 1 study (mean age 32.6 years) and 900 participants were enrolled to phase 2 study (mean age 43.5 years), to receive vaccine or placebo with a two-dose or three-dose schedule. For both trials, local and systemic adverse reactions were absent or mild in most participants. There were no serious adverse events related to vaccine in either trial. After three doses, neutralizing antibodies were detected in all participants receiving either 25 μg or 50 μg dose of vaccine in phase 1 study, and in 97% (the 25 μg group) and 93% (the 50 μg group) of participants, respectively, in phase 2 study. The SARS-CoV-2-neutralizing geometric mean titres (GMTs) were 94.5 for the 25 μg group and 117.8 for the 50 μg group in phase 1, and 102.5 for the 25 μg group and 69.1 for the 50 μg group in phase 2, exceeding the level of a panel of COVID-19 convalescent samples (GMT, 51). Vaccine induced balanced T_H_1 and T_H_2 responses. The 50 μg group did not show enhanced immunogenicity compared with the 25 μg group.

**Interpretation:** The protein subunit vaccine ZF2001 is well-tolerated and immunogenic. The safety and immunogenicity data from phase 1 and 2 trials for ZF2001 support the use of 25 μg vaccine dose with three-dose schedule to an ongoing phase 3 large-scale evaluation for safety and efficacy.

**Funding:** National Program on Key Research Project of China, National Science and Technology Major Projects of Drug Discovery, Strategic Priority Research Program of the Chinese Academy of Sciences, and Anhui Zhifei Longcom Biopharmaceutical.

## Research in context

### Evidence before this study

We searched for ClinicalTrials.gov and PubMed up to Dec 11, 2020, with the terms “COVID-19”, “SARS-CoV-2” and “vaccine”. No language restrictions were applied. We identified 56 vaccine candidates being evaluated in clinical trials, with 12 in phase 3 trials. Thirteen phase 1 and/or phase 2 studies have been published, regarding to 4 inactivated vaccines, 3 adenovirus vectored vaccines, 2 mRNA-based vaccines and 1 protein subunit vaccine. Two phase 3 studies have been published. These vaccines with published data are mainly targeting either whole virus or spike protein. One RBD-based vaccine (BNT162b1, developed by BioNTech and Pfizer) was reported using mRNA expressing RBD-trimer, showing the vaccine is tolerant and immunogenic. The only published protein subunit vaccine (NVX-CoV2373, developed by Novavax) is based on modified spike protein with Matrix-M1 as adjuvant. To date, a large number of RBD-based protein subunit vaccines are under clinical and pre-clinical evaluations (http://www.who.int), but no clinical studies have been published.

### Added value of this study

ZF2001 is one of the two protein-based COVID-19 vaccine candidates that advanced into phase 3 clinical trials but is the first RBD-based protein subunit vaccine reported for clinical data. Clinical data for protein subunit COVID-19 vaccine using conventional alum adjuvant was first described here. The phase 1 and phase 2 trials evaluated the safety profile, immunogenicity, dose-effect and vaccination schedule for ZF2001. This candidate is well-tolerated, without vaccine-related serious adverse events. Three immunizations at day 0, 30 and 60 achieved 93-100% seroconversion of neutralizing antibodies, with the GMTs exceeding the magnitude of the convalescent samples. Also, this vaccine elicited moderate cellular immune responses, detected as the balanced production of T_H_1/T_H_2 cell-associated cytokines.

### Implications of all the available evidence

Our findings indicated that RBD-based protein subunit vaccine is safe and immunogenic. Further clinical trials should be performed to investigate its protective efficacy

## Introduction

The coronavirus disease in 2019 (COVID-19) pandemic caused by severe acute respiratory syndrome coronavirus 2 (SARS-CoV-2) infection is still growing worldwide.^1^ As of Dec 11, 2020, over 69 million people have been diagnosed with COVID-19 in 220 countries and regions, with 1.5 million deaths (http://www.who.int). To date, no COVID-19 vaccine has been licensed, although several of them have been approved for emergency use authorization.^2^

Since the COVID-19 pandemic, safe and effective vaccines are highly demanded to control the disease and reboot the society and economics. After one-year global efforts, great progresses have been made for COVID-19 vaccine development,^3-17^ with several candidates developed based on mRNA or virus-vector platform announced protective efficacy more than 70-90% in preventing COVID-19 after an interim analysis.^18-20^ However, these vaccines so far cannot meet the global vaccination demands from different subpopulations living in different surroundings. Besides, the efficacy duration and vaccine-related safety concerns are yet to be further determined.^21^ An ideal COVID-19 vaccine should be of high protective efficacy, but safe and deployable for billions of potential vaccinees. To achieve this goal, optimal vaccine target, platform and regimen are of high importance.^17^ Nowadays, more than 80 vaccines documented in WHO are based on protein subunit platform, suggesting a major platform for COVID-19 vaccine development (http://www.who.int).

To response the pandemic, we launched a vaccine development program against SARS-CoV-2. The resulting vaccine candidate, ZF2001, is one of the two protein subunit vaccines that has advanced into phase 3 trial (NCT04646590). Compared with other vaccine candidates in clinical trials targeting mainly whole virus or full-length spike (S) protein, ZF2001 targets SARS-CoV-2 S protein receptor-binding domain (RBD). RBD is responsible for the engagement of its cellular receptor human angiotensin converting enzyme 2 (hACE2) and is an attractive vaccine target to induce immune responses focusing on blocking the receptor binding.^22-24^ We rationally designed a dimeric form of SARS-CoV-2 RBD with high yields at level of g/L and dramatically enhanced immunogenicity compared with the conventional monomeric form of RBD in animal model.^25^ ZF2001 is formulated by RBD-dimer protein produced in Chinese hamster ovary (CHO) cells adjuvanted with aluminum hydroxide.

We started the phase 1 trial from June 22, 2020 and phase 2 trial from July 12, 2020 in China for the vaccine candidate ZF2001. Here, we report the pooled analysis of both trials, in which, vaccine safety and immunogenicity were assessed. Appropriate vaccine dose and vaccination schedule were determined for efficacy evaluation in phase 3 trial.

## Methods

### Trial design and participants

The phase 1 trial was conducted at two hospitals in China: The Second Affiliated Hospital of Chongqing Medical University (Chongqing) and Beijing Chao-Yang Hospital of Capital Medical University (Beijing). The phase 2 trial was conducted at Hunan provincial center for disease control and prevention (Xiangtan, China). Eligible participants were healthy men and nonpregnant women, 18 to 59 years of age. Healthy status, assessed during the screening period, was based on medical history and clinical laboratory findings, vital signs, and physical examination. Volunteers who had a history of SARS or COVID-19, or were tested positive for SARS-CoV-2 exposure (by real-time PCR assay or enzyme-linked immunosorbent assay [ELISA]), or contacted with confirmed COVID-19 patients were excluded. Exclusion criteria also included a history of seizures or mental illness; allergy to any ingredient in the vaccine; acute febrile disease in 24 hours and digestive diseases in 7 days before enrollment; congenital or acquired immune diseases; serious chronic disease; abnormal chest CT image; positive test of HBV, HCV, HIV and syphilis; tumor patients; receipt of any blood products in the past 3 months; receipt of any research medicines or vaccine in the past 3 month; and being unable to comply with the study schedule. Full details of the eligibility criteria are described in the trial protocol provided in the appendix.

The trial was designed with double-blind, randomized and placebo-parallel-controlled. Participants in phase 1 were divided into 3 groups at a ratio of 2:2:1 and allocated to receive three doses of vaccine (25 μg or 50 μg antigen dose) or placebo on day 0, 30 and 60. Participants in phase 2 were divided into six groups at a 1:1:1:1:1:1 ratio, with the former three groups receiving two doses of vaccine (25 μg or 50 μg antigen dose) or placebo, 30 days apart, while the latter three groups receiving three doses of vaccine (25 μg or 50 μg antigen dose) or placebo, 30 days apart.

Participants were recruited through community recruitment advertisements. All participants provided written informed consent before enrollment in the trial. The trial protocol was approved by the institutional review board of The Second Affiliated Hospital of Chongqing Medical University, Beijing Chao-Yang Hospital of Capital Medical University, Hunan Provincial Center for Disease Control, Prevention and National Medical Products Administration (NMPA), China and was performed in accordance with the Declaration of Helsinki and Good Clinical Practice. Safety oversight for specific vaccination pause rules and for advancement was performed by an independent safety monitoring committee.

### Randomisation and masking

The randomisation statisticians used SAS statistical software (version 9.4) to generate the random table of participants in each dose group. These randomisation statisticians were not allowed to participate in other related work of these clinical trials and not allowed to disclose the blind code to any personnel participating in this clinical trial. The participants were randomly assigned to the experimental vaccine group or the placebo group according to the block randomisation method, with the block and block size of 5 and 5, respectively, in phase 1 trial, and of 75 and 12, respectively, in phase 2 trial. Investigators at trial site assign study numbers strictly according to the order of screening sequence of eligible subjects, and experimental vaccines were obtained and administered according to the numbers. The participants, field investigators and laboratory team were always blinded to group allocation during the trial.

### Procedures

The vaccine was jointly developed by Institute of Microbiology, Chinese Academy of Sciences and Anhui Zhifei Longcom Biopharmaceutical Co., Ltd. ^25^ The vaccine was manufactured according to current Good Manufacturing Practice by Zhifei Longcom Biopharmaceutical Co., Ltd. The recombined vaccine encodes the SARS-CoV-2 RBD antigen (residues 319-537) in dimeric form and was manufactured as a liquid formulation containing 25 μg or 50 μg per 0.5 mL in a vial, with aluminum hydroxide as the adjuvant. Vaccines were stored at 2 °C to 8 °C before use. Vaccine or placebo was administrated intramuscularly in the arm of each participant.

For both trials, participants were observed in the observation room for 30-60 minutes after each dose of vaccination for emerging adverse events. During 0-7 days after each vaccination, any adverse events were self-reported by participants daily on the diary cards, but verified by investigators. The adverse events during 8-30 days after each vaccination were reported by participants through contact cards. All serious adverse events within 1 year after the first dose of vaccination were reported. Solicited local adverse reactions at injection site within 7 days post-vaccination included pain, swelling, induration, redness, rash and pruritus and solicited systemic adverse reactions within 7 days post-vaccination included fever, cough, dyspnea, diarrhea, anorexia, nausea, vomiting, muscle pain (non-vaccination site), arthritis, joint pain, headache, fatigue, acute allergic reaction, irritation or inhibition and mental disorder. Laboratory safety tests including routine blood, and serum chemistry and routine urine were to assess any toxic effects post-vaccination. Adverse events and abnormal changes in laboratory tests were graded according to the latest scale issued by China NMPA (Version 2019).

The sera were collected to evaluate the binding responses against RBD antigen and neutralizing activity against live SARS-CoV-2. Peripheral blood mononuclear cells (PBMC) were collected to assess the specific T cell responses. Blood samples were taken from participants at the scheduled site visits before the vaccination, on day 14, 30, 37, 60, 67, 90 since first vaccination in phase 1, and on day 14, 30, 44, 60, 74, 90 in phase 2. We used ELISA kits (Wantai BioPharm, Beijing, China) to evaluated the binding antibody responses against the RBD proteins according to the manufacturer’s protocol. We also assessed the neutralizing activities against live SARS-CoV-2 by microcytopathogenic effect assay (See Appendix supplementary Methods). PBMCs from whole blood were isolated and stored in liquid nitrogen in advance, then thawed before testing. An enzyme-linked immunospot (ELISpot) assay was used to quantify the specific T-cell responses as previously reported method (See Appendix supplementary Methods). PBMCs were stimulated with overlapping peptide pools covering RBD protein for about 20-24 hours before detection, and the number of spot-forming cells per 1,000,000 cells were calculated. We measured the cytokine secretion of IFN-γ, IL-2, IL-4 and IL-5 with BD™ ELISPOT.

A panel of convalescent serum samples with PCR-positive SARS-CoV-2 infection were obtained from symptomatic hospitalized patients, outpatients and those with asymptomatic infection detected by health-care worker surveillance. The neutralizing activities of these serum samples against live SARS-CoV-2 were measured by microcytopathogenic effect assay.

### Outcomes

For phase 1 trial, primary outcome was the safety of the COVID-19 vaccine. Secondary outcome was the immunogenicity. For phase 2 trial, primary outcomes were both safety and immunogenicity.

The outcomes for safety were occurrence of the adverse events between first injection to 30 days after final injection, including all adverse events, adverse events related to vaccination, adverse events at grade 3 and above, adverse events lead to participants exit. It also included all serious adverse events occurred between first injection to 1 year after final vaccination. All adverse events and serious adverse events related to vaccination were analyzed. Comparison between groups with different doses and dose regimens were analyzed.

The outcomes for immunogenicity were the seroconversion rate and magnitude of the RBD-binding antibody, SARS-CoV-2 neutralizing antibody and T-cell cytokine production. Groups with different doses and schedules were compared and analyzed. Serum samples from vaccine group and the convalescents were compared to assess and interpret the immunogenicity of the vaccine.

### Statistical analysis

Based on clinical consideration and recommendation of pilot trial and technical guideline issued by the NMPA in China, the sample sizes for two trials were practical, and were not determined on a formal statistical power calculation. We firstly compiled statistics of the number and ratio of participants with adverse reactions or serious adverse reactions post-vaccination and compared safety difference among the dose groups using Fisher’s exact probability method. The RBD-binding antibodies, neutralizing titers against live virus were calculated with positive proportion, geometric mean titres (GMTs). The cytokine production was calculated with GMTs. We used the Clopper-Pearson method to calculate 95% confidence intervals (CIs), χ^2^ test/Fisher exact probability method to compare the proportion differences among the dose groups and Student’s t-test to analyze significance of GMTs between different time points. Statistical analyses were using SAS (version 9.4) and GraphPad Prism (version 8.0.1).

### Role of the funding source

The funders of the study had no role in data collection, data analysis, data interpretation, or writing of the Article. All authors had full access to all the data in the study and had final responsibility for the decision to submit for publication.

## Results

In phase 1, a total of 107 volunteers were recruited and screened for eligibility between June 22 and July 3, 2020 in Chongqing and Beijing, China (figure 1). 57 individuals were excluded, leaving 50 eligible participants in the trial. They were randomly divided into 3 groups to receive vaccine (25 μg [n=20] or 50 μg [n=20] RBD-dimer, with adjuvant) or placebo (adjuvant-only [n=10]). The mean age of all participants was 32.6 years (SD 9.41; range 20.9-57.9), with balanced age and sex distribution among vaccination groups (table 1). 8 out of 10 participants in placebo group, 15 out of 20 participants in the 25 μg group and 18 out of 20 participants in 50 μg group completed three-dose immunization and follow-up visits as schedule (figure 1, appendix 1).

**Table 1:** Baseline demographic characteristics of participants in phase 1 and phase 2 trials.

|  | Cohort in phase 1 |  |  | Cohort in phase 2 |  |  |  |  |  |
| --- | --- | --- | --- | --- | --- | --- | --- | --- | --- |
|  | Placebo,<br>3-dose<br>(n=10) | 25 µg,<br>3-dose<br>(n=20) | 50 µg,<br>3-dose<br>(n=20) | Placebo,<br>2-dose<br>(n=150) | 25 µg,<br>2-dose<br>(n=150) | 50 µg,<br>2-dose<br>(n=150) | Placebo,<br>3-dose<br>(n=150) | 25 µg, 3-dose<br>(n=150) | 50 µg,<br>3-dose<br>(n=150) |
| **Ages,**<br>**(year)** |  |  |  |  |  |  |  |  |  |
| Mean<br>(SD) | 32.5<br>(11.3) | 31.7<br>(9.7) | 33.6<br>(8.5) | 44.1<br>(9.3) | 43.04<br>(9.6) | 44.4<br>(8.4) | 43.4<br>(8.9) | 42.7<br>(9.4) | 43.2<br>(9.4) |
| Median | 29.4 | 26.7 | 34.1 | 46.1 | 43.7 | 45.3 | 44.4 | 43.7 | 43.7 |
| Min, Max | 21.0,<br>57.9 | 22.9,<br>54.7 | 20.9,<br>49.4 | 20.7,<br>59.4 | 18.8,<br>58.4 | 19.9,<br>59.1 | 20.7,<br>58.0 | 20.0,<br>59.7 | 19.3,<br>59.6 |
| **Ethnic**<br>**group** |  |  |  |  |  |  |  |  |  |
| Chinese<br>Han<br>nationality | 9 (90%) | 20<br>(100%) | 18<br>(90%) | 149<br>(99%) | 150<br>(100%) | 150<br>(100%) | 149<br>(99%) | 150<br>(100%) | 148<br>(99%) |
| Others | 1 (10%) | 0 | 2<br>(10%) | 1 (1%) | 0 | 0 | 1 (1%) | 0 | 2 (1%) |
| **Sex** |  |  |  |  |  |  |  |  |  |
| Male | 5 (50%) | 14<br>(70%) | 11<br>(55%) | 68 (45%) | 65<br>(43%) | 57<br>(38%) | 74<br>(49%) | 71 (47%) | 63<br>(42%) |
| Female | 5 (50%) | 6 (30%) | 9<br>(45%) | 82 (55%) | 85<br>(57%) | 93<br>(62%) | 76<br>(51%) | 79 (53%) | 87<br>(58%) |

Data are mean (SD) or n (%)

**Figure 1:**
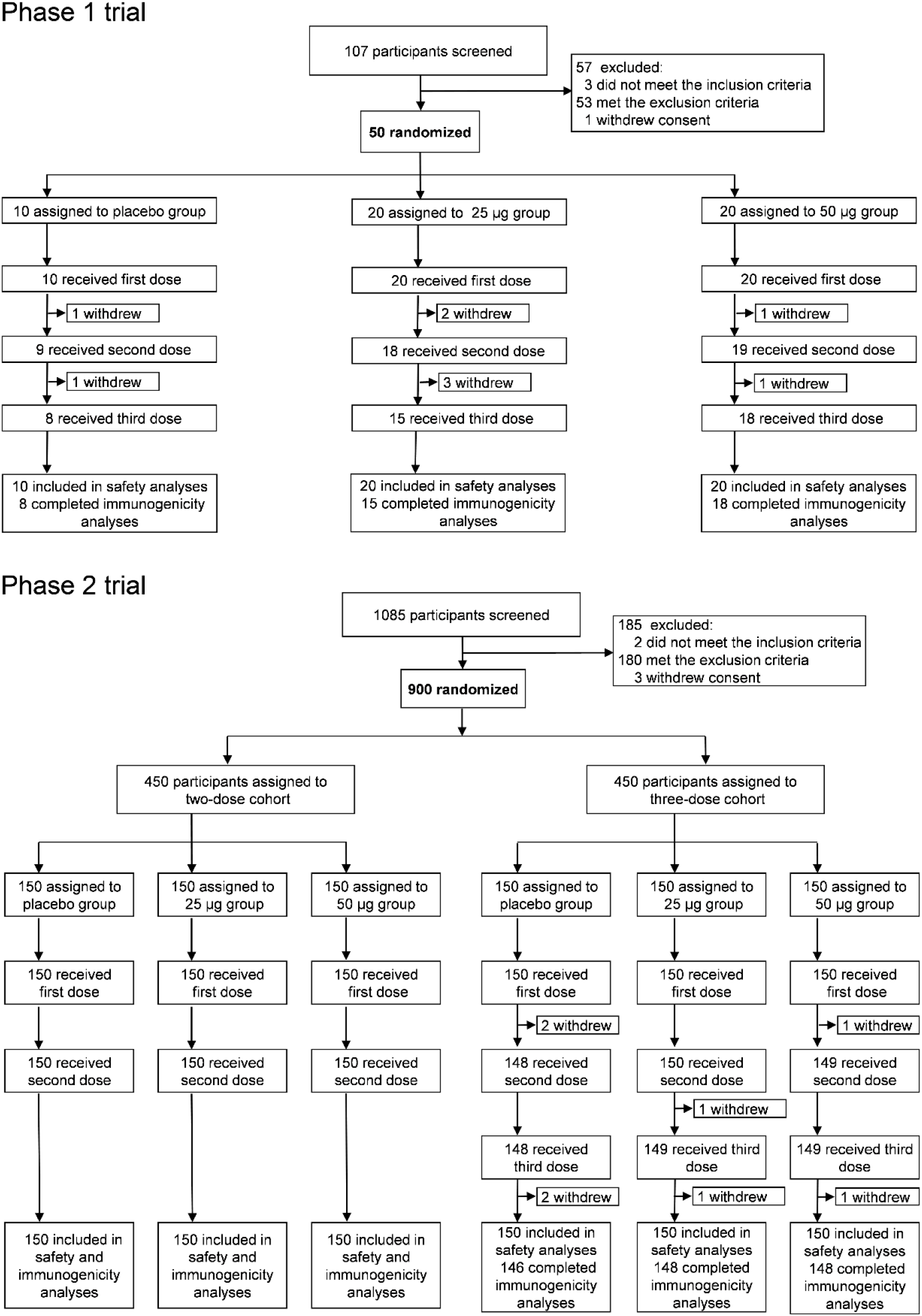
Study profile for the phase 1 and phase 2 trials. The reasons why 17 participants (9 in phase 1, and 8 in phase 2) were not included in the according-to-protocol cohort for vaccination and immunogenicity analysis are listed in the appendix 1.

In phase 2, a total of 1085 volunteers were recruited and screened for eligibility between July 12 and July 17, 2020, in Xiangtan, China (figure 1). 185 volunteers were excluded, and 900 volunteers were enrolled to participant the trial. They were randomly assigned to 6 groups (n=150) to receive vaccine (25 μg or 50 μg RBD-dimer, with adjuvant) or placebo (adjuvant-only) with either a two-dose or a three-dose schedule, 30 days apart. The mean age of the participants was 43.5 years (SD 9.17; range 18.8-59.7) with balanced age and sex distribution among vaccination groups (table 1). All 450 participants with two-dose schedule completed full vaccination and the follow-up visits. For participants with three-dose schedule, 146 out of 150 participants in placebo group, 148 out of 150 participants in the 25 μg group and 148 out of 150 participants in the 50 μg group received full vaccination and completed the follow-up visits (figure 1, appendix 1).

For the overall adverse events within 30 days post-vaccination in phase 1 trial, 6 (60%) of 10 participants in the placebo group, 14 (70%) of 20 participants in the 25 μg group and 18 (90%) of 20 participants in the 50 μg group reported at least one adverse event (figure 2, appendix 2). No statistical significance between these three groups (P=0.1786). Within 7 days post each vaccination, most of the local and systemic reactogenicity was absent or mild (grade 1 or 2 adverse events). The most common solicited local adverse reactions in the placebo, the 25 μg and 50 μg groups were injection-site pain, reported by 20%, 20% and 55%; redness, reported by 10%, 20% and 20%; and itch, reported by 0%, 20% and 35%. The most common solicited systemic adverse reactions in placebo, the 25 μg and 50 μg groups were cough, reported by 0%, 5% and 15%; fever, reported by 0%, 10% and 0%; and headache, reported by 0%, 5%, 5% (figure 2, appendix 2). Two grade 3 (or above) adverse events (10%) were reported in the 50 μg group. One was vaccine-related (swelling and redness) and the other one was serious adverse event (rhabdomyolysis), but was vaccine-unrelated as assessed by the investigators (appendix 9). No vaccine-related serious adverse events or adverse events of special interest were reported.

**Figure 2:**
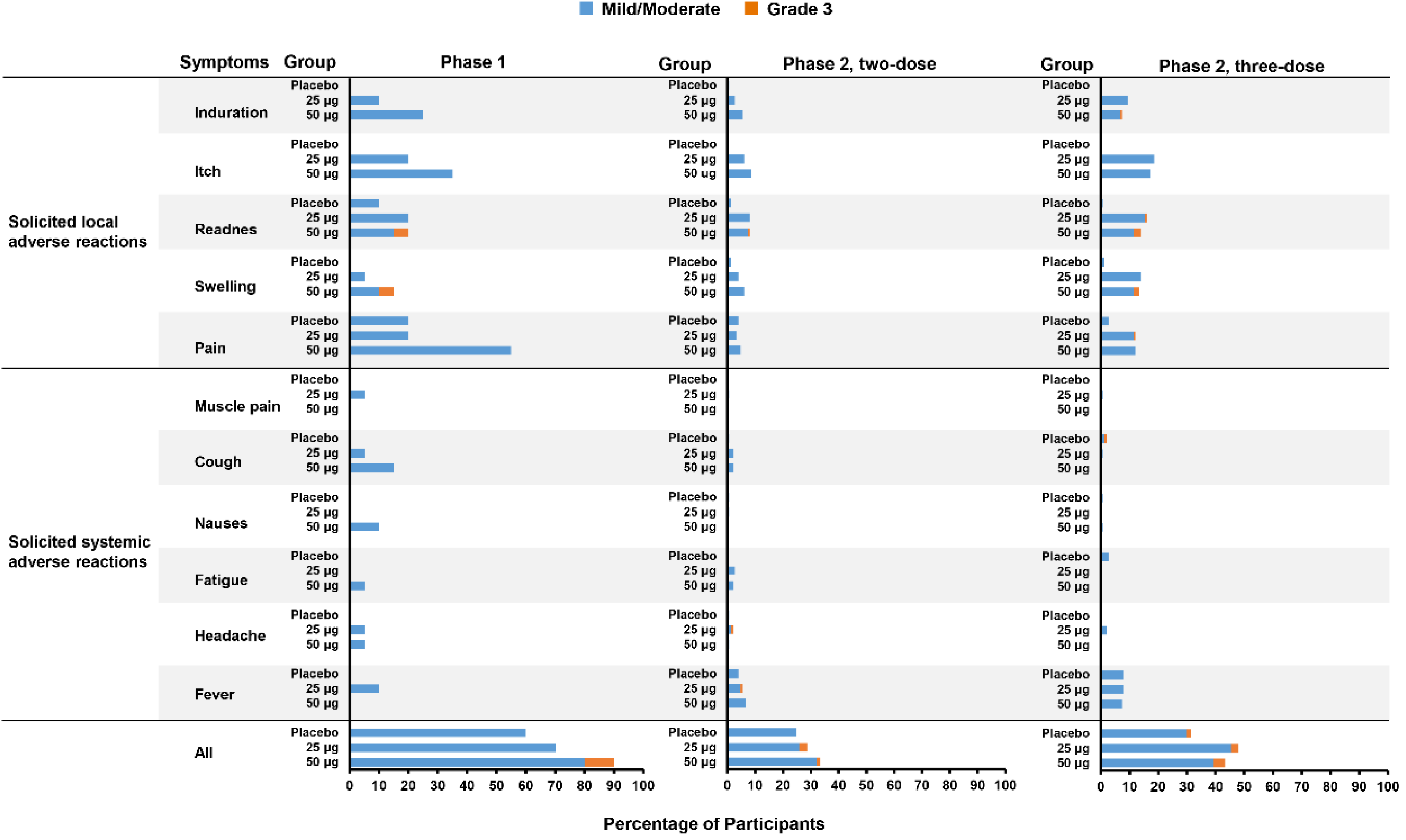
Solicited and unsolicited adverse events in phase 1 and 2 trials. Adverse events within 7 days post each vaccination and overall adverse events within 30 days post full vaccination are shown as percentage of participants in each group. Adverse reactions and events were graded according to the scale issued by National Medical Products Administration, China. The mild and moderate adverse events are colored in blue. The grade 3 or above (severe) adverse events are colored in orange.

In phase 2 trial, the overall adverse events were low within 30 days after vaccination. For participants receiving two-dose vaccinations, 37 (25%) in placebo group, 43 (29%) in the 25 μg group and 50 (33%) in the 50 μg group reported at least one adverse event. For participants receiving three-dose vaccinations, 47 (31%) in placebo group, 72 (48%) in the 25 μg group and 65 (43%) in the 50 μg group reported at least one adverse event (figure 2, appendix 3). Within 7 days after each vaccination, most of the local and systemic reactogenicity was absent or mild (grade 1 or 2 adverse events). The most common solicited local adverse reactions in placebo, the 25 μg and 50 μg groups were injection-site pain, reported by 4%, 3% and 5% in two-dose participants, 3%, 12% and 12% in three-dose participants; swelling, reported by 1%, 4% and 6% in two-dose participants, 1%, 14% and 13% in three-dose participants; induration, reported by 1%, 3% and 5% in two-dose participants, 0%, 9% and 7% in three-dose participants; redness, reported by 1%, 8% and 8% in two-dose participants, 1%, 16% and 14% in three-dose participants; and itch, reported by 0%, 6% and 9% in two-dose participants, 0%, 19% and 17% in three-dose participants. The most common solicited systemic adverse reactions in the placebo, the 25 μg and 50 μg groups were fever, reported by 4%, 5% and 7% in two-dose participants, 8%, 8% and 7% in three-dose participants; cough, reported by 1%, 2% and 2% in two-dose participants, 1%, 1% and 0% in three-dose participants; and headache, reported by reported by 1%, 2% and 1% in two-dose participants, 0%, 2% and 0% in three-dose participants; and fatigue, reported by 0%, 3% and 2% in two-dose participants, 3%, 0% and 0% in three-dose participants (figure 2, appendix 3). 18 participants were reported grade 3 (or above) adverse events, with 12 of them (1.3%) were vaccine-related, including redness (5 cases), swelling (3 cases), injection-site pain (1 cases), induration (1 case), headache (1 case) and cough (1 case). Seven participants were reported serious adverse events, but none of them were considered to related to the study vaccine as assessed by the investigators (appendix 9). Besides, no adverse events of special interest were reported.

Regarding to the immunogenicity, in phase 1 trial we detected the serologic RBD-binding IgG titers by ELISA assay to assess the antibody responses. The baseline antibody titres of the participants are shown in the appendix 4. At day 30 after the first immunization, the seroconversion rates were 61% for the 25 μg group and 79% for the 50 μg group; and were increased to 100% for both groups at day 30 after the second and third immunizations (figure 3A; appendix 5). At day 30 after the first dose, the GMTs of vaccine-induced RBD-binding IgG were 23.1 (95% CI, 11.2-47.9) and 40.8 (95% CI, 22.5-74.0) for the 25 μg and 50 μg group, respectively; and were increased to 1077.0 (95% CI, 663.7-1747.5) and 825.5 (95% CI, 486.9-1399.4) at day 30 after the second dose; and were further enhanced to 2719.5 (95% CI, 1584.0-4668.8) and 2776.8 (95% CI, 1875.5-4111.2) at day 30 after the third dose (figure 3B; appendix 5). The GMTs were markedly increased after the second and third doses in both groups. To analyze the neutralizing antibody titres against live SARS-CoV-2, we measured the 50% neutralizing titres for the serum samples of the participants. The baseline titres of neutralizing antibodies of the participants are given in the appendix 4. At day 30 after the second dose, seroconversion rates reached 93.3% in the 25 μg group and 94.4% in the 50 μg group. At day 7 after the third immunization, the seroconversion rates were 100% in both groups (figure 3C; appendix 6). The SARS-CoV-2-neutralizing GMTs were 14.0 (95% CI, 8.0-24.6) and 11.4 (95% CI, 6.6-19.8) in the 25 μg and 50 μg group, respectively, at day 7 after the second dose; and further enhanced to 94.5 (95% CI, 49.3-181.3) and 117.8 (95% CI, 64.6-214.9) in the 25 μg and 50 μg group, respectively, at day 7 after the third dose (figure 3D; appendix 6). The neutralizing titres dramatically increased after the second and the third immunizations in both groups. Meanwhile, a panel of 89 COVID-19 human convalescent serum samples was tested for comparison. The GMT of neutralizing antibodies for the COVID-19 convalescent serum panel was 51 (95% CI, 38.3-70.5). Encouragingly, the neutralizing GMTs for both the 25 μg and 50 μg groups were approximately two times of the magnitude than human convalescent samples (figure 3D).

**Figure 3:**
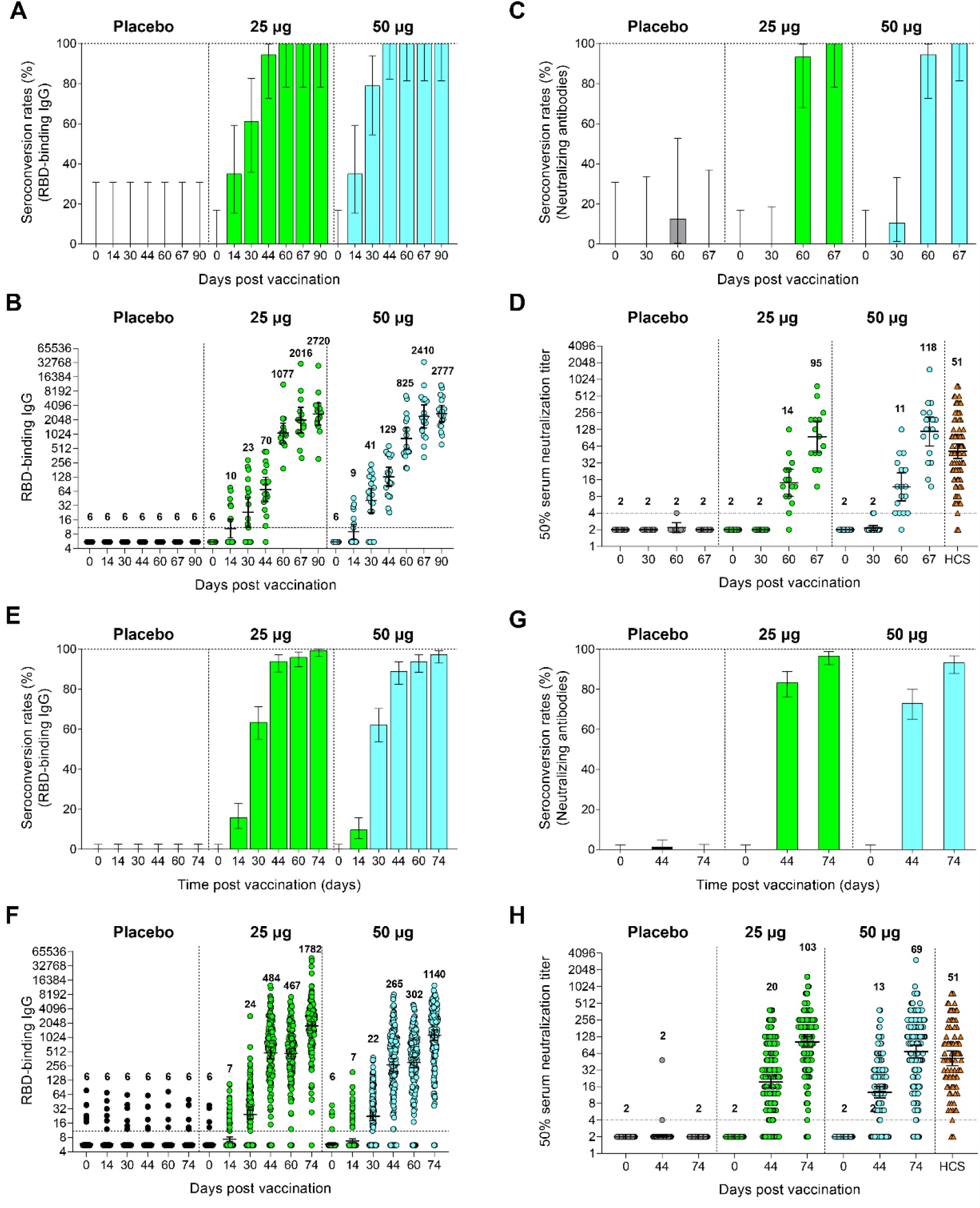
Humoral immune responses in phase 1 and phase 2 trials. Shown are humoral responses in phase 1 trial (A-D) and phase 2 trial (E-H). Seroconversion rates (A and E) and geometric mean titres (GMTs) (B and F) of RBD-binding antibody at different time points after vaccination are shown. Seroconversion rates (C and G) and GMTs (D and H) of neutralizing antibody at different time points after vaccination are shown. Error bars show 95% confidence intervals (CIs). The horizontal dashed line indicated the limit of detection. HCS = human convalescent serum.

In phase 2 trial, the baseline antibody titres of the participants are shown in the appendix 4. At day 30 after the first immunization, the seroconversion rates were 59% and 65% for the 25 μg and 50 μg group, respectively, in two-dose participants; 63% and 62% for the 25 μg and 50 μg group, respectively, in three-dose participants. At day 30 after the second immunization, the seroconversion rates were increased to 95% and 97% for the 25 μg and 50 μg group, respectively, in two-dose participants; 96% and 94% for the 25 μg and 50 μg, respectively, in three-dose participants. At day 14 after the third immunization, the seroconversion rates were further increased to 99% and 97% for the 25 μg and 50 μg group, respectively, in three-dose participants (figure 3E; appendix 7). In participants with two-dose schedule, RBD-binding IgG GMTs significantly increased from 19.4 (95% CI, 16.0-23.6) (30 days after the 1^st^ immunization) to 419.5 (95% CI, 325.8-540.1) (30 days after the 2^nd^ immunization) for the 25 μg group; and from 22.6 (95% CI, 18.5-27.6) (30 days after the 1^st^ immunization) to 344.8 (95% CI, 271.0-438.7) (30 days after the 2^nd^ immunization) for the the 50 μg group (appendix 7). In participants with three-dose schedule, RBD-binding IgG GMTs significantly increased after each immunization, from 24.4 (95% CI, 19.6-30.2) (30 days after the 1^st^ immunization), to 467.3 (95% CI, 366.8-595.3) (30 days after the 2^nd^ immunization), and to 1782.3 (95% CI, 1440.2-2205.7) (14 days after the 3^rd^ immunization), for the 25 μg group; and from 22.2 (95% CI, 18.1-27.4) (30 days after 1^st^ immunization), to 302.5 (95% CI, 234.2-390.6) (30 days after 2^nd^ immunization), to 1140.0 (95% CI, 882.2-1473.2) (14 days after the 3^rd^ immunization), for the 50 μg group (figure 3F; appendix 7). Regarding to the neutralizing antibody titres against live SARS-CoV-2, seroconversion rates and GMTs were analyzed. For the participants with two-dose schedule, 76% and 72% were seroconverted in the 25 μg and 50 μg group, respectively, at 14 days after the 2^nd^ immunization (appendix 8). For the participants with three-dose schedule, 83% and 73% were seroconverted at 14 days after the 2^nd^ immunization in the 25 μg and 50 μg group, respectively; and further increased to 97% and 93%, respectively, at 14 days after the 3^rd^ immunization (figure 3G, appendix 8). The neutralizing GMT against live SARS-CoV-2 in participants with three-dose schedule was boosted from 19.5 (95% CI, 15.2-25.0) (14 days after 2^nd^ immunization) to 102.5 (95% CI, 81.8-128.5) (14 days after 3^rd^ immunization) in the 25 μg group; and from 12.6 (95% CI, 10.0-16.0) (14 days after 2^nd^ immunization) to 69.1 (95% CI, 53.0-90.0) (14 days after 3^rd^ immunization) in the 50 μg group (figure 3H, appendix 8). After three-dose vaccination, both the 25 μg and 50 μg groups showed neutralizing GMTs exceeding the level of a panel of convalescent sera (figure 3H). Again, there is no enhancement of neutralizing GMT for the participants receiving high vaccine dose (50 μg) compared with those receiving low vaccine dose (25 μg).

To assess the T-cell responses, we performed ELISpot assay for the peripheral blood mononuclear cells (PBMCs) of participants in phase 1 trial against SARS-CoV-2 RBD protein. T_H_1 and T_H_2 cell responses were measured by detection of the T_H_1 cytokines IFN-γ, IL-2 and the T_H_2 cytokines IL-4, IL-5. As expected, placebo group did not show obvious increase for all tested T-cell cytokines. By contrast, either the 25 μg or 50 μg group elicited moderate levels of both T_H_1 (IFN-γ and IL-2) and T_H_2 (IL-4 and IL-5) cytokine production post vaccinations (figure 4). Therefore, ZF2001 induced balanced T_H_1/T_H_2 responses in humans.

**Figure 4:**
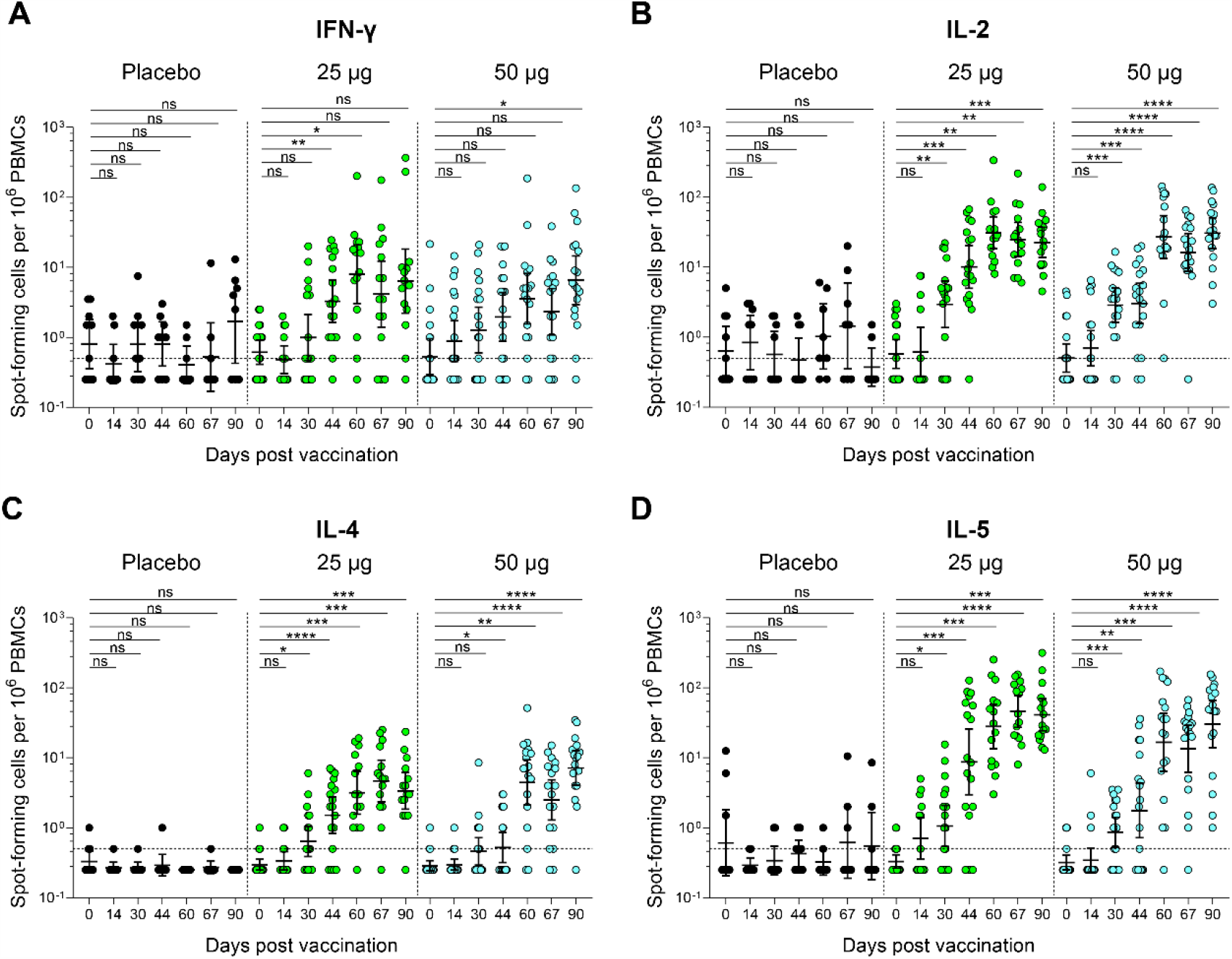

## Discussion

S protein RBD is an attractive vaccine target against coronavirus.^26^ To date, a number of vaccine candidates based on RBD have been demonstrated efficient in animal models against SARS-CoV, MERS-CoV or SARS-CoV-2.^17^ Several RBD-based COVID-19 vaccines are being evaluated in clinical trials.^17^ Clinical data have been published for an RBD-based COVID-19 vaccine candidate, developed by BioNTech/Pfizer. This candidate, BNT162b1, is based on mRNA strategy and showed good immunogenicity in humans.^14,15^ Here, we reported the first clinical study for the RBD-based protein subunit vaccine against coronavirus in a population of 950 health adult participants.

We found vaccination with different doses (25 or 50 μg) and schedules (two- or three-doses) were well-tolerated. The incidents of adverse reactions between vaccine groups and placebo groups were similar in both phase 1 and 2 trials, without statistical significance. Most of the adverse reactions were mild, with the most common symptoms being injection-pain, redness and swelling. These adverse reactions were anticipated for alum-adjuvanted protein subunit vaccines and were transient and resolved within 3-4 days post-vaccination. Compared with other COVID-19 vaccine candidates, such as mRNA vaccines or adenovirus-vectored vaccines, the occurrences of fever and fatigue were lower.^3-7,10,11,13^ Compared with another protein subunit vaccine, NVX-CoV2373, using Matrix-M1 as adjuvant, the occurrences of injection-site pain, fatigue, headache and nausea were lower, as well.^12^

Humoral responses have been considered as immune correlates of protection against SARS-CoV-2.^27-31^ So far, humoral responses for several COVID-19 vaccine candidates in clinical trials have been reported. It should be noted that currently it is difficult to compare different vaccine candidates as there are no standardized neutralization assays. We used microcytopathic effect assay to determine neutralizing antibody titres for the serum samples from vaccinees, with a panel of COVID-19 convalescent sera tested as the control. The comparative immunogenicity showed three-dose of ZF2001 elicited neutralizing GMT two times greater than the GMT tested for the convalescent sera. Notably, two mRNA-based vaccines and two adenovirus-vectored vaccines have been announced 70-90% efficacy in preventing COVID-19 in phase 3 clinical trials, with their vaccine-induced humoral responses being reported similar or higher than those from the convalescent samples.^3-5,13,18^ Although the interpretation of the vaccine immunogenicity can be influenced by the illness severity of the convalescent selected for comparison, our study indicated encouragingly that ZF2001 has the potential to elicit protective humoral responses against COVID-19. Three-dose vaccination was found dramatically enhanced the antibody responses compared with two-dose vaccination, however, increasing the antigen dose from 25 μg to 50 μg did not show an improvement for the immunogenicity. Therefore, vaccine of 25 μg antigen with three-dose schedule was determined for efficacy evaluation in phase 3 trial.

Cellular immune responses also play protective roles in SARS-CoV-2 infections.^32,33^ Virus-specific T-cell responses were associated with milder disease in COVID-19 patients.^34^ Based on the levels of spot-forming cells per million cells, cytokines representing both T_H_1 and T_H_2 responses were moderately elicited post vaccination. The moderated but balanced production of T_H_1 and T_H_2 cell-associated cytokines may benefit for the T-cell-mediated protection rather than the potential risk of vaccine-enhanced respiratory disease which is presumably caused by predominant T_H_2-biased responses.^35^

This trial report has several limitations. First, the participants in both trials were young adults aged from 18-59, not including juvenile and elderly persons, whereas, the elderly is more vulnerable to SARS-CoV-2. Moreover, the participants had limited ethnic diversity, mainly the ethnic Han. These limitations will be addressed by our outreach study for cohort covering broader age range and ethnic backgrounds. Second, the immunogenicity was tested at day 30 post full vaccination in phase 1 and day 14 post full vaccination in phase 2, which cannot be assessed for the duration of the immune response. Immune responses at later timepoints including those at least 6 months post vaccination will be obtained in the follow-up visits and investigation. Third, there was still no benchmark to evaluate protective immune responses against COVID-19. Therefore, although ZF2001 induced neutralizing GMTs higher than the convalescent samples, it is yet hard to predict its protective efficacy. Since no participant had SARS-CoV-2 exposure, we were unable to assess the protective efficacy and vaccine-associated enhanced diseases at this moment. This limitation will be addressed in the ongoing international multi-center phase 3 trials.

## Data Availability

Individual participant data will be made available on request, directed to the corresponding author (GFG). After approval of a proposal, data can be shared through a secure online platform.

## Contributors

GFG, LD and JY initiated and designed the vaccine. SY, YL (Yan Liu) and JY contributed to the protocol and design of the trials. XY, HR and LL were the study site principal investigators in phase 1 trial. FL was the study site principal investigators in phase 2 trial. LG and JL were responsible for the organization and supervision of the phase 2 trial. ZZ participated in the implementation of trials. YL (Yan Li), ZH, JY, JW, PH, CL, SM, XZ, XF, CW, XD, ST, FW and LW were contributed to the laboratory testing and assay development. ZC was responsible for the collection of convalescent serum samples. EH and CW were responsible for vaccine manufacture. YL (Yi Luo) did the statistical analysis. SY, LD, JY and GFG participated in the data collection and analysis for manuscript. LD, YL (Yan Li) and GFG contributed to the writing of the manuscript.

## Declaration of interests

SY, EH, CW, and ZZ are employees of Anhui Zhifei Longcom Biopharmaceutical Co. Ltd. YL (Yan Liu) is an employee of Chongqing Medleader Bio-Pharm Co. Ltd. YL (Yi Luo) is an employee of Beijing Keytech Statistical Technology Co. Ltd. LD, YL, JY and GFG are listed as inventors on pending patent applications for RBD-dimer-based CoV vaccines. The pending patents for RBD-dimers as protein subunit vaccines for SARS-CoV-2 have been licensed to Anhui Zhifei Longcom Biopharmaceutical Co. Ltd, China.

## Acknowledgments

This work is funded by National Program on Key Research Project of China (2020YFC0842300), National Science and Technology Major Projects of Drug Discovery (2018ZX09101001), Strategic Priority Research Program of the Chinese Academy of Sciences (XDB29010202) and Anhui Zhifei Longcom Biopharmaceutical. We thank Fengcai Zhu, Hudachuan Jiang and Pengfei Jin from Jiangsu Provincial Center of Disease Control and Prevention (Jiangsu, China) for their valuable help for the manuscript preparation. We thank Yuhai Bi for providing the virus and the staff of Biosafety Level 3 Laboratory for their help in live virus experiments (Institute of Microbiology, Chinese Academy of Science) We thank Zhifang Ying and Zhen Chen for their technical support in neutralization experiments (National Institute for Food and Drug Control)

## Notes

### Clinical Trial

NCT04445194, NCT04466085

### Author Declarations

The trial protocol was approved by the institutional review board of The Second Affiliated Hospital of Chongqing Medical University, Beijing Chao-Yang Hospital of Capital Medical University, Hunan Provincial Center for Disease Control, Prevention and National Medical Products Administration (NMPA), China and was performed in accordance with the Declaration of Helsinki and Good Clinical Practice.

